# Evaluating Knowledge Fusion Models on Detecting Adverse Drug Events in Text

**DOI:** 10.1101/2024.02.14.24302829

**Authors:** Philipp Wegner, Holger Fröhlich, Sumit Madan

## Abstract

**Background:** Detecting adverse drug events (ADE) of drugs that are already available on the market is an essential part of the pharmacovigilance work conducted by both medical regulatory bodies and the pharmaceutical industry. Concerns regarding drug safety and economic interests serve as motivating factors for the efforts to identify ADEs. Hereby, social media platforms play an important role as a valuable source of reports on ADEs, particularly through collecting posts discussing adverse events associated with specific drugs.

**Methodology:** We aim with our study to assess the effectiveness of knowledge fusion approaches in combination with transformer-based NLP models to extract ADE mentions from diverse datasets, for instance, texts from Twitter, websites like askapatient.com, and drug labels. The extraction task is formulated as a named entity recognition (NER) problem. The proposed methodology involves applying fusion learning methods to enhance the performance of transformer-based language models with additional contextual knowledge from ontologies or knowledge graphs. Additionally, the study introduces a multi-modal architecture that combines transformer-based language models with graph attention networks (GAT) to identify ADE spans in textual data.

**Results:** A multi-modality model consisting of the ERNIE model with knowledge on drugs reached an F_1_-score of 71.84% on CADEC corpus. Additionally, a combination of a graph attention network with BERT resulted in an F_1_-score of 65.16% on SMM4H corpus. Impressively, the same model achieved an F_1_-score of 72.50% on the PSYTAR corpus, 79.54% on the ADE corpus, and 94.15% on the TAC corpus. Except for the CADEC corpus, the knowledge fusion models consistently outperformed the baseline model, BERT.

**Conclusion:** Our study demonstrates the significance of context knowledge in improving the performance of knowledge fusion models for detecting ADEs from various types of textual data.

**Author Summary:** Adverse Drug Events (ADEs) are one of the main aspects of drug safety and play an important role during all phases of drug development, including post-marketing pharmacovigilance. Negative experiences with medications are frequently reported in textual form by individuals themselves through official reporting systems or social media posts, as well as by doctors in their medical notes. Automated extraction of ADEs allows us to identify these in large amounts of text as they are produced every day on various platforms. The text sources vary highly in structure and the type of language included which imposes certain challenges on extraction systems. This work investigates to which extent knowledge fusion models may overcome these challenges by fusing structured knowledge coming from ontologies with language models such as BERT. This is of great interest since the scientific community provides highly curated resources in the form of ontologies that can be utilized for tasks such as extracting ADEs from texts.

## Introduction

An adverse drug event (ADE) can be defined as “an injury resulting from a medical intervention related to a drug” [1]. ADEs as a major aspect of drug safety are objective of interest in the pharmacovigilance efforts done by pharmacological companies as well as medical regulatory bodies. Negative experiences with certain medications are frequently reported in textual form by individuals themselves through official reporting systems or social media posts, as well as by doctors in their medical notes. The mentioned ADEs are often hidden in unstructured text, and the process of identifying and extraction of ADE entities from such text requires a significant amount of a medical professional’s time. Performing large-scale automatic extraction from a variety of text sources could help domain experts in quickly identifying new ADEs. However, this extraction process requires robust and highly accurate text mining methods.

In recent years, the natural language processing (NLP) field has made significant advancements with transformer-based language models such as BERT [2] or GPT [3]. These models have set new benchmarks in several NLP tasks. Furthermore, these models have been successfully applied to detect ADEs from textual documents [1, 4–6]. There are mainly two different types of texts mentioning ADEs such as reports or scientific publications written by medical professionals and reports provided by the patient or their relatives themselves. Social media texts differ from medical reports as they often contain informal language, slang, abbreviations, and colloquialisms. Additionally, these texts predominantly consist of opinions of people and contain fewer factual statements. Due to the continuously growing quantity and significance of social media texts,we place particular attention on analyzing patient-reported texts. In this work, we considered the CADEC corpus [5] that contains annotated texts from https://askapatient.com, which is a forum dedicated to collecting drug experiences and a corpus, here referred to as SMM4H [6], that comprises annotated Twitter postings. Moreover, we evaluate our models on three more corpora, namely PSYTAR [7], TAC [8], and ADE [9]. The CADEC, SMM4H, and PSYTAR were derived from sources where patients authored the texts themselves, whereas the ADE and TAC were composed by medical experts written in formal and scientific language. Further details on the corpora are given in Section Datasets.

It is important to highlight previous scientific initiatives that have aimed to extract ADEs from texts. Sboev et al. [10] elaborated on the performance of various transformer models evaluated on CADEC, where they reported an F_1_-score of 69.68% for strict matches (exact matching between true and predicted instances) using the XLM-Roberta-large model that ranked best among all considered models. Additionally, Portelli et al. [11] provided a performance overview of different transformer models on CADEC and SMM4H, in which they reported F_1_-scores of 67.95% and 62.15%, respectively. Portelli et al. [11] reported that a SpanBERT-based approach yielded the best results. Furthermore, Ge et al. [4] offered a federated learning methodology for the ADE detection problem and evaluated it on both datasets. This approach was able to achieve for relaxed matches (partial overlap of true and predicted instances) an F_1_ of 84.55% on CADEC and 67.8% on SMM4H corpus. For strict matches, 65.16% and 32.69% were reported for the same corpora by the authors. Ramesh et al. [12] presented their solution to the 2021 SMM4H shared task 1 that adopts the roBERTa base model to extract ADE mentions, which reached a relaxed F_1_-score of 50% on the final test set. Furthermore, Raval et al. [13] presented an interesting strategy by tackling text classification concerning ADEs as well as the actual ADE span extraction with a multi-task learning approach that used the T5 as a pre-trained encoder-decoder transformer model. They could reach the strict F_1_-score of 69.8% on CADEC and 71.3% on SMM4H corpus as well as the relaxed F_1_-scores of 79.1% and 75.1%, respectively. Another notable work that deserves mention is of Haq et al. [14] as they evaluated their NLP pipeline on the ADE corpus [9] as well as on CADEC and SMM4H. The end-to-end system proposed by Haq et al. was able to report strict macro-averaged F_1_-scores of 91.7%, 78.7%, and 76.7% on the ADE, CADEC, and SMM4H corpora respectively. Furthermore, Miftahutdinov & Tutubalina [15] evaluated BERT on the PSYTAR corpus and were able to reach an accuracy of 83.07% during the task of normalizing the ADE entities to a controlled vocabulary. Analogously the authors reported accuracy scores of 88.84% on CADEC as well as 89.64% on SMM4H during the entity normalization task. Finally, in the 2017 Text Analysis Conference (TAC) a team from the University of Texas Health Science Center at Houston was able to achieve a micro-averaged F_1_-score of 82.48% over all entities of the TAC corpus including ADE mentions. The participants from Houston were able to reach that score by utilizing a bi-directional LSTM model.

Moreover, Stanovsky et al. [16] adopted a fusion learning approach by combining contextual knowledge from DBpedia with a Bi-LSTM. By doing so the authors reported an F1-score of 93.4% on the CADEC corpus. Fusion model approaches are often able to increase performance in comparison with standalone transformer models. Zhang et al. [17] reported a performance increase from 73.5% F_1_-score using a BERT model to 75.5% adopting ERNIE as a fusion learning model evaluating however on the Open Entity dataset [18]. Liu et al. [19] published an alternative approach that demonstrates the advantages of transformer-based language encoding with contextual knowledge, Their K-BERT model achieved a notable increase of 0.04 in the F1-score on a question-answering task.

In this study, we conducted a series of experiments to assess the effectiveness of knowledge fusion methods in combination with transformer-based NLP models for extracting ADEs from unstructured texts. We performed these experiments on a total of five diverse text corpora. To incorporate contextualized knowledge, we constructed a knowledge graph (KG) that included drug brand names and integrated a symptom ontology. This combination proved to be well-suited for analyzing ADE-related texts. Additionally, we utilized graph neural network (GNN) techniques, specifically a graph attention network, to learn representations of drug and symptom entities within the KG. These representations were subsequently integrated into transformer models through a fusion learning approach. We compared our proposed model architecture against ERNIE, a well-established knowledge fusion language model, as well as two non-knowledge fusion models, namely BERT and BioBERT.

## Materials and Methodology

First, we introduce different datasets and knowledge resources used in our work and subsequently we present the knowledge fusion models that have been developed for the purpose of detecting ADEs from textual corpora.

### Datasets

#### CADEC

The CSIRO Adverse Drug Event Corpus (CADEC) [5] is an annotated text corpus published in 2015 that consists of forum posts from askapatient.com and comes with 5 different types of annotations: ADE, Drug, Disease, Symptom, and Finding (any other clinical finding).

The whole CADEC corpus includes reports on 12 drugs such as Diclofenac or Lipitor. Diclofenac (https://go.drugbank.com/drugs/DB00586) is a non-steroidal anti-inflammatory drug that is used to treat pain and inflammation from different sources while Lipitor (https://go.drugbank.com/drugs/DB01076) lowers lipid levels and reduces the risk of cardiovascular diseases. The CADEC corpus is composed of 1,253 posts with 7,398 sentences in total, where 1,107 posts contain at least one ADE mention (see Table 1). This adds up to 7,409 ADE spans with an average post length of six sentences. Finally, all posts were written between January 2001 and September 2013 by patients between 17 and 84.

**Table 1.** Overview of the ADE datasets used in this study. Note that the SMM4H corpus does not contain any drug annotations.

| Dataset | Document class | # Documents | # Sentences | # Drugs | # ADEs |
| --- | --- | --- | --- | --- | --- |
| CADEC [5] | Drug reviews | 1,253 | 7,398 | 1,800 | 7,409 |
| SMM4H Subtask 1b [6] | Tweets | 1,300 | 2,107 | - | 1,496 |
| PsyTAR [7] | Drug reviews | 891 | 6,009 | 792 | 4,813 |
| TAC Task 1 [8] | Drug labels | 101 | 3,154 | 249 | 13,795 |
| ADE [9] | Medline case reports | 2,972 | 4,272 | 5,063 | 5,776 |

#### SMM4H

The second dataset used in this work is the SMM4H corpus [6], which is one of the datasets provided to the participants of the Social Media Mining for Health Applications (#SMM4H) Shared Task 2021 (https://healthlanguageprocessing.org/smm4h-shared-task-2021/). In this work, we focus on the corpus for Subtask 1b, which is about extracting ADE mentions from Twitter posts. We ignore Subtasks 1a which dealt with classifying Tweets containing an ADE and 1c which tackled the normalization of ADEs to MedDRA.

There are differences between the SMM4H Subtask 1b corpus and the CADEC, while the biggest difference might be that CADEC has annotations of 5 different types whereas the corpus of Subtask 1b of SMM4H has only adverse drug reaction mentions tagged. The corpus is composed of 1,300 tweets with 1,800 annotated ADE spans (see Table 1). On average each tweet has 21 words and two sentences.

#### PsyTAR

The third corpus considered in this work is the corpus presented by Zolnoori et al. [7]. The “Psychiatric Treatment Adverse Reactions” (PsyTAR) corpus contains 891 drug reviews from askapatient.com which is the same source as the previously mentioned CADEC corpus. The corpus contains reviews for four drugs (Zoloft, Lexapro, Cymbalta, and Effexor XR) and holds a total of 6009 sentences with 4813 ADE mentions (see Table 1). On average each post contains 7 (6.7) sentences. Further note that the PsyTAR text corpus contains, besides ADE mentions, 6 other annotation types, which are Withdrawal Symptoms (WDs), Signs/Symptoms/Illness (SSIs), Drug Indications (DIs), Drug Effectiveness (EF), and Drug Infectiveness (INF) and other, not applicable, mentions.

#### TAC

The TAC corpus [8] was assembled from drug labels and was used in the 2017 text annotation conference (TAC). The corpus consists of a set of drug labels in which ADE mentions among

other entities are annotated. In that conference participants were provided with the corpus and challenged to extract adverse drug reactions from these drug labels. This task was referred to as Task 1 within TAC. Each drug label contains on average 79 sentences and hence was split into sentences to fit it into the transformer models used in this work. Each sentence contains on average 33 (32.69) words. Besides ADE entities the corpus comes with annotations for Severity, Factor (additional aspects of the ADE entity), Drug Class, Negation, and Animal.

#### ADE

The 5th and final corpus used was published by Gurulingappa et al. [9] and was constructed from 3000 MEDLINE case reports. After an exhaustive annotation and harmonization process that involved three annotators, the corpus holds 2972 reports. The final corpus comprises a total of 5063 drugs and 5776 ADE annotations distributed over 4272 sentences (see Table 1). On average each sentence contains 20 (20.09) words. Besides drug and ADE entities the corpus further contains annotations for Dosage. Other than some of the corpora previously introduced, the authors of the ADE corpus did not restrict the retrieved documents to a certain set of drugs but rather retrieved 30.000 documents and randomly selected the 3000 case reports that were further used for the annotation process.

### Knowledge Bases

In our work, we explored the enhancement of transformer models by incorporating contextual knowledge through fusion models to improve the detection of adverse drug events. We utilized two knowledge resources: one for encoding knowledge about symptoms and the other for modeling the domain of drug space.

### Symptom Ontology

The symptom ontology (SYMP) is a publicly available ontology developed in the context of the Gemina system [20]. The creators designed the ontology while understanding a symptom as a “perceived change in function, sensation or appearance reported by a patient indicative of a disease” [20]. The ontology consists of 860 classes as well as a total of 1,586 cross-references to other databases like UMLS (https://www.nlm.nih.gov/research/umls/index.html) or ICD (https://www.who.int/standards/classifications/classification-of-diseases). Furthermore, the ontology comprises 5,445 axioms and class annotations such as definitions, synonyms, and labels of symptoms. We use the symptoms ontology to provide context knowledge about symptoms. An example of how a model can enrich sentences with symptom classes is shown in Fig. 1.

**Fig. 1.**
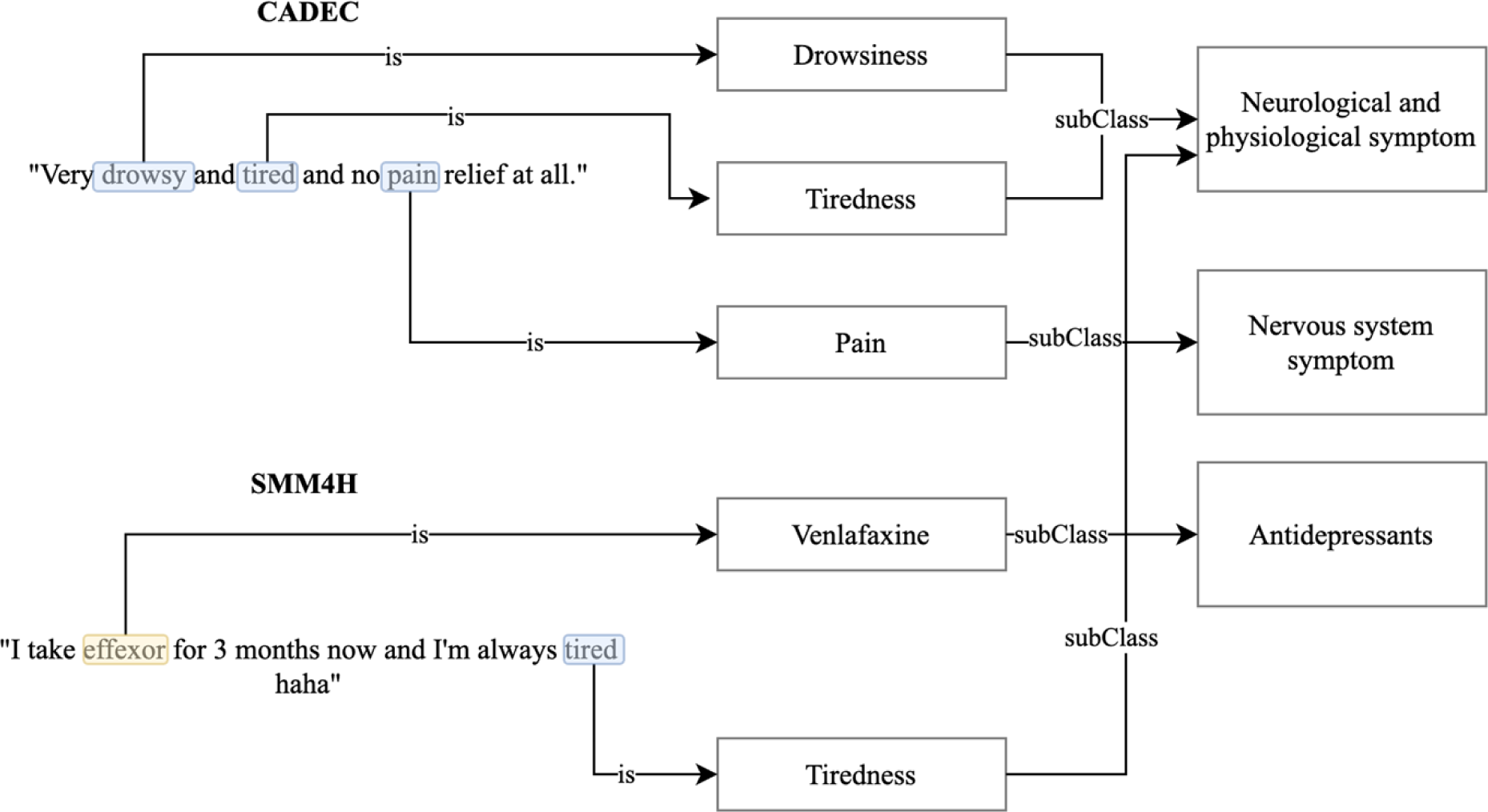
CADEC and SMM4H example phrases that are enriched with contextual knowledge about drugs and symptoms. The sentence from CADEC “Very drowsy and tired and no pain relief at all.” can be equipped with symptom classes such as Drowsiness and Tiredness, which are subclasses of Neurological and physiological symptom class, as well as Pain, which is a subclass of Nervous system symptom.

### Drug Resources

Contextual knowledge about drugs and how they function in the human body can be valuable for tackling the task of ADE detection. We decided to assemble such knowledge in a structured way and store it in the form of an ontology. The resulting ontology inherits information from the ATC ontology and is further enriched with selected information about drugs. Fig. 1 illustrates an example of how a model can enhance sentences by incorporating drug resource information. Fig. 1 depicts the utilization of contextual knowledge exemplarily for CADEC and SMM4H but works equally for the other three corpora.

We used three different resources to collect various information on approved drugs. Firstly, the DrugBank database (version 5.1.9) [21] was used to extract drug descriptions, synonyms, and product names, as well as information about drug targets. Fortunately, DrugBank provides cross-references to the anatomical therapeutic chemical classification system (ATC), which divides active ingredients into classes based on anatomical properties like the organ they act on, chemical properties, as well as therapeutic properties [22]. DrugMechDB [23] is another drug resource, which contains information about the mechanism of action of a drug in the body. This mechanism is represented as a graph where each node can be of several types (such as disease, drug, protein, or cell). A sub-graph was taken from this graph to obtain information about the proteins that are involved in the drug mechanism, which we added to our ontology. Furthermore, since this ontology is used to extract drug entities from text based on the drug product names it is important to add as many brand names to the ontology as known. To accomplish that, the website drugs.com was a highly useful resource for adding brand names for each drug in ATC.

Finally, all of the collected knowledge on drugs was added to the ATC ontology at its respective position and stored as an OWL (web ontology language) file. The resulting ontology, in this work referred to as DRUGO, provides knowledge about drug names, definitions, synonyms, drug targets, and information about proteins involved in the drug’s action mechanism. The final DRUGO ontology comprises a total of 6,441 classes.

### Detection of Adverse Drug Events

Our experimental strategy to create models that can detect ADEs in texts builds upon knowledge fusion models that integrate transformer-based models with knowledge graph embeddings. As transformer-based models, we focus on using BERT [24] and BioBERT [25]. These models are also used to create baseline results. Furthermore, we experiment with multiple fusion approaches such as ERNIE and the graph concat model, which are introduced in the next sections.

### Knowledge Fusion

To incorporate the information from the aforementioned knowledge bases (DRUGO and SYMP) into the language models, a numerical representation is necessary that effectively captures the encoded knowledge. We experimented with two approaches, the first one uses the well-established TransE method [26] to embed the underlying graphs of the two ontologies into a vector space. Whereas, in the second approach, a GNN was incorporated for this task. More specifically a graph attention network (GAT) was trained with a node classification task, which provided the final node-level embeddings for the integration in the language model.

A total of three GATs were trained on the DRUGO and SYMP ontologies, as well as on an ontology generated by combining SYMP and DRUGO. In this approach, ontologies are treated as graphs, without taking into account any logical axioms, similar to other ontology embedding approaches. All GATs have been trained identically by initially considering the ontologies as graphs and assembling a set of nodes (V) from the classes of the ontology and a set of edges (E) from the relations between the classes. Specifically, we derived E by treating every ‘subClass’ property as an edge. As a result, we obtained a circle-free, fully connected, directed graph with 6,441 nodes and 6,440 edges for DRUGO, 860 nodes and 859 edges for SYMP, and, 7301 nodes and 7300 edges for the combined KG of DRUGO and SYMP.

In the following step, initial representations for all nodes were generated. This was performed by using the annotation properties of each ontology class/node and embedding these using a pre-trained language model. For all graphs, this was done by using either BERT or BioBERT, depending on the exact experimental setup. This lead to the representation of each node as a 768-dimensional real vector. Graphs derived from DRUGO and SYMP provided a top-level classification with 14 classes, enabling the assignment of each node to one of these classes based on its position in the graph. The third graph obtained from combining the two ontologies yielded 28 classes.

Finally, a GNN was trained to predict the assigned class of each node in the graph. Note that in our work, we specifically favored GAT over other GNN architectures because of its capability for self-attention. The self-attention mechanism in GAT allows nodes to attend to the features of their neighboring nodes. With the usage of GAT, we would like to address the issue that certain classes of the ontology may lack valuable information due to a lack of class annotations. As a result, nodes can assign lower weights to neighbors without valuable information due to the attention mechanism [27]. The aforementioned methodology of generating knowledge graph embeddings corresponds to what Yang et al. [28] refer to as cascaded model architecture. In this architecture, initial node features are generated using language models and then further processed by GNNs [28].

### Integrating Transformer-based Models with GNNs

We propose a knowledge fusion model to combine node embeddings learned via a graph neural network with a transformer-based model. We begin by taking an input sentence and using a rule-based tagger to identify symptoms and/or drug entities depending on the given knowledge graph. The KG can be either SYMP, DRUGO, or a combination of both. The tagged input sequence has the same length as the original input sequence but holds additional information for those input tokens that were tagged by the rule-based annotator. Furtheron, the tagged input sequence is passed through a GNN and returns a vector that holds zeroes for tokens that do not belong to any tagged entity and the corresponding node embedding for tokens that were tagged by the previous tagger. Subsequently, the resulting vector *v* is aligned with the representation of the transformer (by adding zeros wherever a padding token was added or where words were split into word pieces). This aligned vector *ṽ* is then concatenated with *T* to create a final knowledge-enriched representation *T̃* of the input sequence. This final representation is further passed into a linear layer, which serves as the classification head (Fig. 2).

**Fig. 2.**
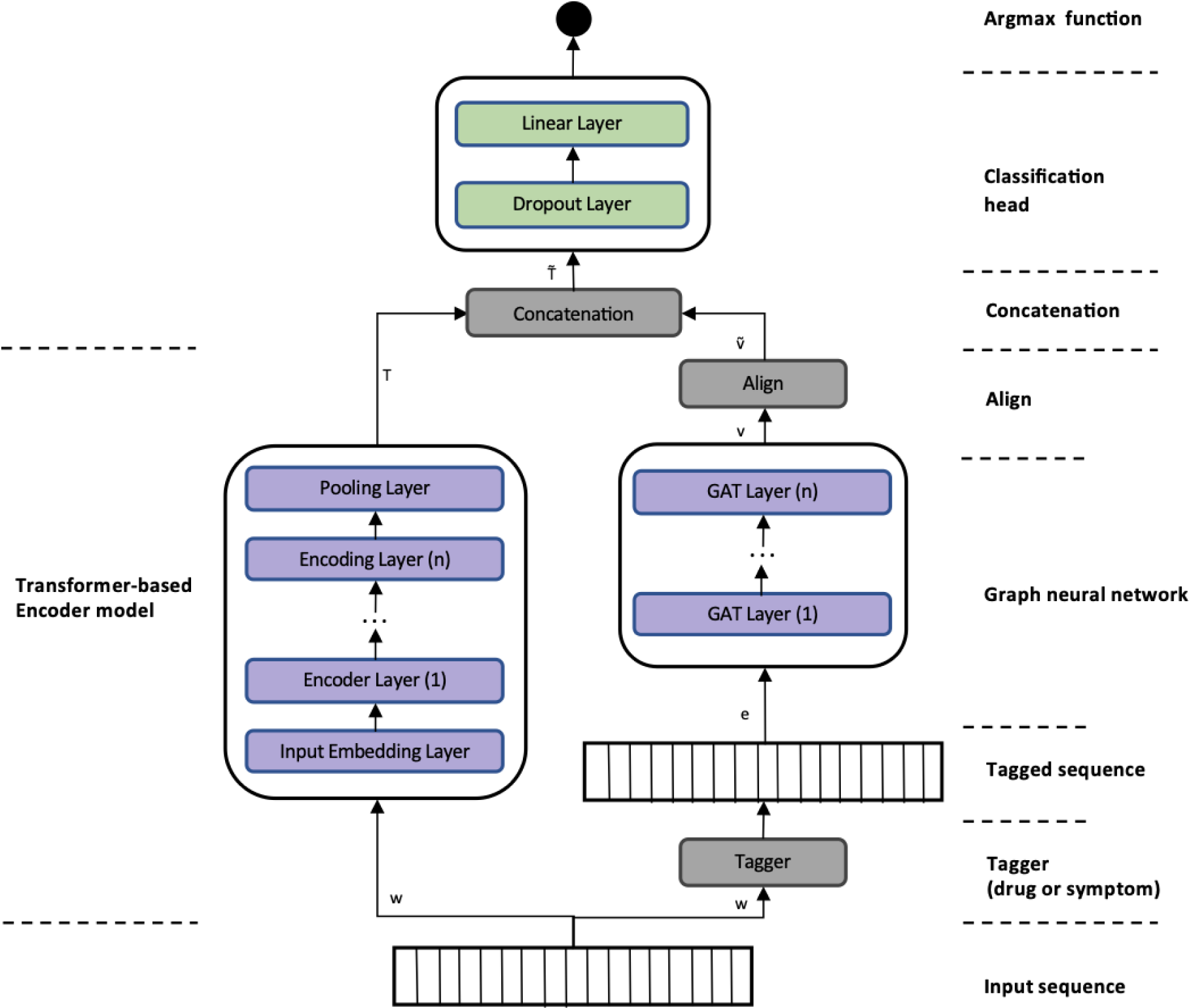
The architecture of the graph concat model with fixed and trainable GNN weights.

Additionally, we set the GNN weights as fixed by default, resulting in the usage of GNN as a lookup table within the underlying embedding space. We refer to this architecture as a graph concat model. Nevertheless, we have implemented an additional model variant called the graph concat adaptive weights model, in which we treat the GNN weights as trainable parameters that are adjusted during the training of the entire model. Fig. 2 illustrates the architecture of the graph concat model without (orange box) and with (purple box) adaptive GNN weights.

Furthermore, instead of using the entire graph as in the setting presented above, we explored an additional GNN configuration where only a subgraph of the knowledge graph is used and passed through the GNN. This subgraph is constructed from the k-hop neighborhood of the tagged entity. Finally, instead of concatenating the node representation to the transformer representation, a graph pooling layer (concatenation of global max and average pooling) is added and its output is concatenated to the transformer representations. The just presented architecture will be noted as graph concat (graph concat AW for adaptive GNN weights and graph concat AWS for graph concat with adaptive weights and subgraph modification) from now on.

### Compared Method: ERNIE

Enhanced Language Representation with Informative Entities (ERNIE) is a fusion model introduced by Zhang et al. [17]. However, ERNIE handles the knowledge injection differently than other models. Instead of calculating the representation of context knowledge within ERNIE itself, it is computed separately. In ERNIE, TransE is utilized to generate and retrieve embeddings for the knowledge. To have a fair comparison, we also adopted this approach in our work. For a more detailed explanation of the working principle of ERNIE, we refer to the original study published by Zhang et al. [17].

The implementation used in this work is obtained from the GitHub repository https://github.com/thunlp/ERNIE, which provides a pre-trained ERNIE model.

### Experimental Setup and Training Strategy

To perform an unbiased final evaluation on a completely independent test set, we randomly chose and reserved 20% from each dataset. The remaining 80% of each dataset was divided into a train and validation set, with a ratio of 4-to-1. This means that 64% of the entire dataset served as a training set used to train the model, while the remaining 16% was used as the validation set hyperparameter tuning. After hyperparameter tuning, we trained the final model by combining both training and validation sets, which were used to evaluate the performance of the aforementioned independent test set. Furthermore, to have maximum comparability along all the different model architectures, those splits were consistently applied throughout all experiments.

Each experiment conducted in our study was constructed from the four categories listed in Table 2. The categories encompass the model architecture, the pre-trained transformer-based language model, the ADE text corpus, and the contextual knowledge resource. The selected model architectures further categorize the results into ERNIE, graph concat model with fixed GNN weights, graph concat model with adaptive GNN weights, and graph concat model with adaptive GNN weights and k-hop subgraph. Additionally, baseline experiments are considered as a separate category that only uses the pre-trained language models BERT and BioBERT. It is important to note that we utilized BERT as a general language model to assess the performance achievable by a transformer-based encoder that was not pre-trained on domain-specific documents. On the other hand, BioBERT is a domain-specific model that was pre-trained on biomedical documents [25]. All models were evaluated on all five ADE text corpora. Finally, each model was equipped with either contextual knowledge about drugs, symptoms, or both. In addition to the 10 baseline experiments, the various options for experiment configurations resulted in a total of 115 experiments.

**Table 2:** Overview of experiment categories. Their combination results in a total of 115 experiments in addition to 10 baseline experiments.

| Experiment categories | Values |
| --- | --- |
| Knowledge fusion model architecture | ERNIE (not used in combination with pre-trained transformer),<br>Graph concat model with fixed GNN weights,<br>Graph concat model with adaptive GNN weights, and<br>Graph concat model with adaptive GNN weights and k-hop subgraph |
| Pre-trained language model | BERT and BioBERT |
| ADE corpora | SMM4H, CADEC, PSYTAR, TAC, and ADE |
| Knowledge resource | SYMP, DRUGO, and DRUGO + SYMP |

To ensure unbiased and comparable results, the same overall strategy for training, validation, hyperparameter tuning, and testing was employed in each experiment. The optimal hyperparameters were deduced by performing Bayesian hyperparameter optimization [29]. To determine the optimal hyperparameters for each model, multiple models with different hyperparameter configurations were trained on the training set. These models were then evaluated on the validation set, to maximize the F_1_-Score. The cross-entropy loss function was employed for all models in the context of NER. The AdamW [30] optimization algorithm was chosen to adjust the model’s weights during training. Finally, the optimal hyperparameters were used to train models on the combination of training and validation sets. These new models were then tested on held-out, independent test sets of each corpora.

### Evaluation Scheme

We used the precision, recall, and F_1_ measures to assess the performance of models. Each dataset was labeled in the IOB scheme with which each token of a sequence is labeled either as outside (O) of a named entity, as the beginning (B), or as an inside (I) token of a named entity. Hence, the classification head of each of the models had three output neurons and the NER problem was formulated as a classification task with three classes. However, we are interested in ADE spans that can consist of multiple tokens, therefore, for the final evaluation the IOB labeling was discarded, and the sequences were aggregated into real ADE mentions. The final scores were then calculated by taking into account the exact overlap of the full spans of ADE mentions.

### Implementation

The experiments conducted in this study were implemented using PyTorch and PyTorch Lightning. An essential component are transformer-based models for which we used the Huggingface transformers library. To perform hyperparameter tuning Optuna was chosen as the library. Finally, for processing and handling the considered datasets we used Pandas and Spacy. The baseline models as well as the graph concat model experiments are using BERT and BioBERT, which come in different sizes and configurations. We used uncased BERT, commonly known as ‘bert-base-uncased’, which contains a total of 110M parameters. The BioBERT model is specified as ‘dmis-lab/biobert-v1.1’, which has the equivalent amount of parameters as ‘bert-base-uncased’. The model training and testing was performed using Nvidia’s V100 and A100 GPUs.

## Results

We evaluated the aforementioned five different model architectures^1^ on each of the ADE datasets. able 3 provides an overview of the final evaluation results providing the F1-score obtained by pplying the models within a certain configuration on the independent test sets. Here the onfiguration refers to the choice of context knowledge resource and underlying transformer-based odel, where applicable. Please take note that the graph concat k-hop subgraph experiments ere omitted from Table 1 since this architecture did not achieve the top ranking on any of the orpora. For a comprehensive overview of results including this architecture as well as precision nd recall measures for all models, we refer to Supplementary Table 1.

When examining the results on the CADEC corpus (Table 3), one may observe that the best baseline experiment utilizing BERT already demonstrates a strong performance in terms of the F_1_-score (71.84%). None of the other models evaluated on CADEC were able to improve upon this score. However, ERNIE equipped with contextual knowledge about drugs achieved the same score of 71.84%. Additionally, the graph concat AW model incorporated with drugs and symptom knowledge came quite close with a score of 71.82%.

**Table 3:** Final evaluation results on test set from all experiments. F_1_ stands for F_1_-score. All scores are strict scores and given in %. The best score on each corpus is given in bold. AW=adaptive weights.

| Model | Knowledge resource | F<sub>1</sub> (in %) on ADE Corpora |  |  |  |  |
| --- | --- | --- | --- | --- | --- | --- |
|  |  | CADEC | SMM4H | PSYTAR | ADE | TAC |
| BERT | - | **71.84** | 62.30 | 70.02 | 75.37 | 92.06 |
| BioBERT | - | 70.81 | 61.95 | 68.80 | 79.42 | 93.87 |
| ERNIE + TransE | DRUG | **71.84** | 63.23 | 70.63 | 75.44 | 92.57 |
| ERNIE + TransE | DRUGO_SYMP | 69.32 | 61.76 | 70.95 | 76.04 | 92.55 |
| ERNIE + TransE | SYMP | 68.70 | 61.32 | 71.40 | 76.58 | 92.06 |
| Graph concat + BERT | DRUG | 70.45 | 62.65 | 71.38 | **79.79** | 93.80 |
| Graph concat + BERT | DRUGO_SYMP | 70.70 | **65.16** | 72.32 | 78.84 | 93.49 |
| Graph concat + BERT | SYMP | 71.05 | 62.83 | 72.03 | 78.13 | 93.87 |
| Graph concat + BioBERT | DRUG | 70.28 | 61.51 | 68.24 | 76.73 | **94.15** |
| Graph concat + BioBERT | DRUGO_SYMP | 69.57 | 62.75 | 70.05 | 78.90 | 93.88 |
| Graph concat + BioBERT | SYMP | 69.40 | 62.48 | 69.32 | 78.59 | 93.31 |
| Graph concat AW + BERT | DRUG | 70.55 | 63.96 | **72.50** | 79.03 | 93.02 |
| Graph concat AW + BERT | DRUGO_SYMP | 71.82 | 63.99 | 71.38 | 79.54 | 93.87 |
| Graph concat AW + BERT | SYMP | 70.59 | 64.22 | 72.02 | 78.4 | 93.22 |
| Graph concat AW + BioBERT | DRUG | 71.23 | 61.05 | 70.08 | 78.11 | 93.75 |
| Graph concat AW + BioBERT | DRUGO_SYMP | 68.87 | 62.12 | 69.62 | 78.62 | 93.78 |
| Graph concat AW + BioBERT | SYMP | 70.16 | 57.78 | 69.00 | 76.01 | 93.45 |

<sup>1</sup> Baseline, ERNIE, Graph concat, Graph concat AW, Graph concat AWSUB

The performance of the models on the SMM4H corpus, in general, was lower than on all other corpora. The difference of performance can already be observed in the results of the baseline experiments that show a noticeable gap of almost 8-20% points. Furthermore, ERNIE, equipped with prior knowledge about drugs, was able to perform better on SMM4H with an F_1_-score of 63.23% than the best baseline experiment using BERT, which reached an F_1_-score of 62.3%. Moreover, the graph concat AW model with contextual knowledge about symptoms adopting BioBERT as the underlying transformer was also able to report better F_1_-scores (64.22%) than the baseline experiments and better than the best-performing ERNIE model (Table 3). Finally, the graph concat model with fixed GNN weights using BERT as its underlying pre-trained transformer while equipped with joint prior knowledge about symptoms and drugs reports the overall best score on SMM4H with an F_1_-score of 65.16%.

On PSYTAR, the ERNIE model equipped with prior knowledge about symptoms, reaching an F_1_-score of 71.40%, was able to slightly improve the performance of the BERT baseline experiment that was able to achieve an F_1_-score of 70.02%. The graph concat model using BioBERT and drugs and symptoms knowledge was able to improve this score to 72.32% F_1_- score. The graph concat AW model with BERT and the drug knowledge graph further improves this score to 72.50% F_1_-score.

On the ADE corpus, the ERNIE model was not able to reach the score reported by the best baseline model BioBERT (79.42% F_1_-score). However, the graph concat AW model using BERT and adopting prior knowledge about drugs and symptoms was able to slightly increase this score to 79.54% F_1_-score. The graph concat model with fixed GNN weights while also using BERT as its transformer and equipped with prior knowledge about drugs further improved this score to 79.79% F_1_-score.

Finally, on the TAC corpus, all models considered in the results were able to score F_1_-scores above 90%. The best baseline model, BioBERT, was able to reach an F_1_-score of 93.87%. The ERNIE and the graph concat AW model were not able to outperform the best baseline model. However, the graph concat model with fixed GNN weights using BioBERT as its transformer and equipped with contextual knowledge about drugs was able to increase upon the baseline performance achieving the highest F_1_-score of 94.15% on TAC corpus.

We performed an additional analysis to determine the different attributes of each of the 5 corpora that could shed some light on explaining the modeling performance. Table 4 depicts the results of this corpus analysis comprising three measures. Firstly, the wordpiece diversity, which was assembled by counting how many unique wordpieces could be found in each sentence of a corpus normalized by the total amount of wordpieces in a sentence. The second measure calculates the sentence length on wordpiece level and the number of hits in the DRUGO_SYMP knowledge graph. A hit is defined as an entity in the sentence corresponding to a node in the knowledge graph. All values presented in Table 4 are averaged over all sentences in the corresponding corpus. The CADEC corpus is a clear outlier in terms of the mean number of KG hits, the mean sentence length, and wordpiece diversity. CADEC is the only corpus where we did not observe any advantage of using a knowledge fusion model in terms of F_1_-score.

**Table 4:**
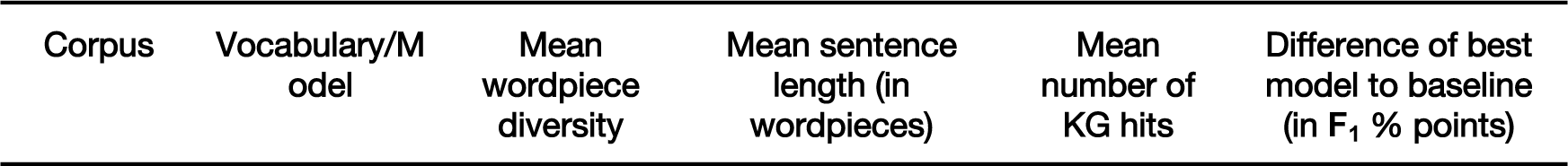
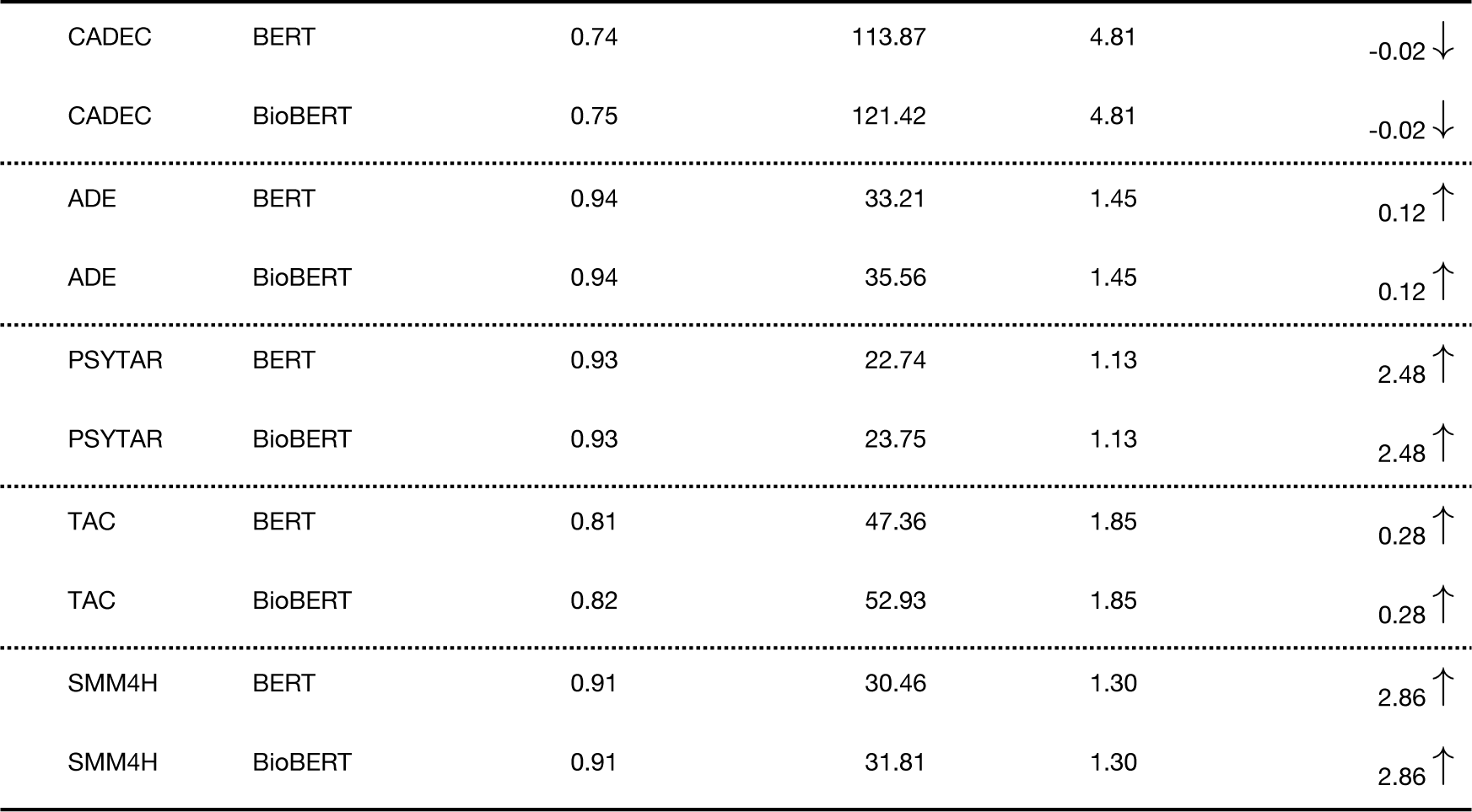
Corpora characterization in terms of average wordpiece diversity, average sentence length, and average number of knowledge graph hits.

| Corpus | Vocabulary/Model | Mean wordpiece diversity | Mean sentence length (in wordpieces) | Mean number of KG hits | Difference of best model to baseline (in $F_1$ % points) |
| --- | --- | --- | --- | --- | --- |
| CADEC | BERT | 0.74 | 113.87 | 4.81 | -0.02 ↓ |
| CADEC | BioBERT | 0.75 | 121.42 | 4.81 | -0.02 ↓ |
| ADE | BERT | 0.94 | 33.21 | 1.45 | 0.12 ↑ |
| ADE | BioBERT | 0.94 | 35.56 | 1.45 | 0.12 ↑ |
| PSYTAR | BERT | 0.93 | 22.74 | 1.13 | 2.48 ↑ |
| PSYTAR | BioBERT | 0.93 | 23.75 | 1.13 | 2.48 ↑ |
| TAC | BERT | 0.81 | 47.36 | 1.85 | 0.28 ↑ |
| TAC | BioBERT | 0.82 | 52.93 | 1.85 | 0.28 ↑ |
| SMM4H | BERT | 0.91 | 30.46 | 1.30 | 2.86 ↑ |
| SMM4H | BioBERT | 0.91 | 31.81 | 1.30 | 2.86 ↑ |

## Discussion

Extracting meaningful insights about ADEs from unstructured text offers the chance to enhance our knowledge of ADEs and in the long run contributes to drug safety. Specifically, the extraction of ADEs from patient-reported texts allows for gathering great amounts of negative drug experiences since vast amounts of data are published every day on social media. In our work, we evaluate various knowledge fusion modeling approaches on the ADE extraction task using five relevant text corpora, namely CADEC, SMM4H, PSYTAR, TAC, and ADE. Additionally, we utilized a rich knowledge base in terms of drugs and symptoms, which provided valuable contextual knowledge to these models. Knowledge graph embeddings derived from GNNs have ensured a knowledge representation well suited for the fusion with linguistic representations obtained using transformer-based large language models. The final results on independent test sets showed that using models with contextual knowledge can help to gain performance on ADE corpora.

We observed a significant variation in performance scores and model behavior across different datasets. There was no clear advantage of adopting a knowledge fusion methodology over the baseline model BERT on the CADEC dataset. Using graph concat adaptive weights model resulted in an F_1_-score quite similar to the BERT and ERNIE model. However, on the SMM4H corpus, we observed a performance increase from top-scoring baseline (BERT) to ERNIE to the graph concat model. BERT reached an F_1_-score of 62.30%, and equipping it with contextual knowledge about drugs and symptoms raised this score to 65.16%. When examining the results for PSYTAR, the top-performing baseline model (BERT) achieved an F_1_-score of 70.02% for extracting ADE entities. ERNIE was able to improve this score by approximately 1.5%. By enabling BERT to utilize contextual knowledge about drugs through the graph concat architecture, the score further increased to 72.5%. When considering the ADE corpus, there was a notable difference in scores between baseline models (75.37% for BERT and 79.42% for BioBERT). None of the ERNIE models were able to match the baseline score achieved by BioBERT. However, the graph concat model with fixed GNN weights that utilizes BERT and contextual knowledge about drugs was able to slightly increase the baseline performance to a 79.79% F_1_-score. Similarly, in the case of the TAC dataset, BioBERT was able to reach a high F_1_-score of 93.87% that was not surpassed by any ERNIE model. The graph concat model was able to slightly increase the baseline performance on TAC to an F_1_-score of 94.15%.

There was no clear indication of whether the graph concat models work better with BERT or BioBERT as the underlying transformer model. However, we observed that on CADEC, utilizing BioBERT in knowledge fusion could improve the baseline BioBERT performance (BioBERT: 70.81% F_1_ and 71.23% F_1_ graph concat with adaptive GNN weights and contextual knowledge about drugs), whereas this could not be observed for BERT (71.84% F_1_ is best score on CADEC). When considering the usefulness of knowledge resources, it is noteworthy to mention that all models that outperformed the baseline experiments relied either on DRUGO or DRUGO_SYMP contextual knowledge. Based on this observation, it suggests that contextual knowledge about drugs may hold greater importance for the knowledge fusion models compared to knowledge about symptoms. The trend was apparent in both the graph concat model and ERNIE.

As mentioned, our observations indicate that the effectiveness of knowledge fusion models varies across different corpora. We did not observe any performance improvement using knowledge fusion models on the CADEC corpus. This aligns with the findings in Table 4, which highlights CADEC being an outlier in the textual analysis in terms of wordpiece diversity, sentence length, and KG hits. Further investigation is necessary to determine the causal relationship between these metrics and the potential improvement of pure linguistic models with knowledge fusion. However, based on our interpretation of the results, it can be reasoned that knowledge fusion models are most beneficial for relatively short text, such as postings found in SMM4H and PSYTAR (<24 wordpieces on average in PSYTAR and <32 in SMM4H). Notably, the CADEC corpus stands out in terms of the number of hits in the knowledge graph. This suggests that an excessive amount of contextual knowledge may not contribute positively to the model’s accuracy. Liu et al. [19] introduced the concept of knowledge noise (KN), which refers to the phenomenon that an excess of context can disrupt the original meaning of the sentence. However, further investigation is needed to find whether during knowledge fusion KN played a role in the lack of performance improvement on CADEC. Additionally, since PSYTAR and SMM4H are derived from Twitter, it is reasonable to assume that these corpora deviate from formal, scientific English. In this context, knowledge fusion can potentially compensate for the informality in language and for the lack of linguistic context by providing valuable information on specific ADEs.

The current workflow infuses context knowledge into models for the words that are identified as drugs or symptoms by a rule-based NER tagger. For this purpose, we preferred a rule-based system to avoid false positives in terms of context knowledge. However, a more advanced machine learning-based tagger with a better performance may produce even higher results, which we will explore in our future work. One possible machine learning-based model for such an approach would be Med7 [31], which reports good results in terms of F_1_-score on the task of extracting drug entities from text. Although the used knowledge resources have shown performance gains while using the knowledge fusion approach, they are far from being complete and perfect. Encoding even more knowledge about drugs and symptoms could improve the current models of ADE detection.

Although this study performed a comprehensive analysis, it is important to note existing limitations. Further knowledge fusion approaches such as K-BERT, K-Adapters, or SKILL [32–34] are worth exploring in future experiments for evaluating knowledge fusion models on the ADE extraction task. Some of the training datasets used in this work comprise only a relatively small number of postings, around 1,000 for both the SMM4H and CADEC corpora. It is well-known that deep learning-based NLP models generally tend to perform better when trained on larger datasets. Therefore, to further enhance the performance of the knowledge fusion models employed in this study, having access to large and diverse corpora of patient-reported texts that include annotated ADE entities, particularly in the style of CADEC, would be beneficial. Consequently, future efforts should be directed toward creating, collecting, and annotating a comprehensive ADE corpus of diverse texts, which could contribute to the advancement of this research.

## Conclusion

The presented work elaborates on the approach to enriching transformer models such as BERT and its relative, BioBERT, with contextual knowledge about the texts fed into them. Two types of prior knowledge on drugs and symptoms were considered in this work. The drug knowledge resource provides rich, structured knowledge about drugs and their working principles and was especially created for this work. We conducted a great number of experiments and reported the combinations of transformer models, knowledge fusion architectures, and context knowledge that yielded the highest F_1_-scores. The presented results allow the conclusion that contextual knowledge encoded suitably and provided to a transformer model is a valid approach to improve performance in an NER task scenario. Also observable is that this prior knowledge is especially of great use when the data at hand is rather unstructured and composed of short texts as is the case in the SMM4H and PSYTAR corpus. Finally, one can conclude that knowledge resources that provide well-structured domain knowledge, encoded as knowledge graphs respectively ontologies can provide valuable context for transformer models. Graph neural networks have shown to be a well-suited method to derive a numerical representation of the ontologies used in this work capable of being concatenated with the linguistic representation created by a transformer model. The architecture of the graph concat model with and without adaptive GNN weights implemented in this work has shown to be advantageous compared to pure transformers (BERT and BioBERT) as well as to another, well-established, knowledge fusion model, ERNIE. Hence, that architecture deserves additional development to further improve its performance on tasks such as ADE extraction in structured and unstructured texts. Huge potential lies in the idea of fusing large language models with appropriate domain knowledge and definitively deserves further research that includes whether the presented approach generalizes on tasks further than detecting adverse drug events in texts.

## Data Availability

The data underlying the results presented in the study are available from various study owners. These studies have been linked in the main manuscript.

## Availability

The code is available in the repository: https://github.com/SCAI-BIO/adr-detection-with-knowledge-fusion.

